# A plus 5-Year Journey of Infective Challenges and Graft outcomes in ABO-Incompatible Kidney Transplants: Insights from a tertiary Care Centre in Saudi Arabia

**DOI:** 10.1101/2025.04.03.25325114

**Authors:** Bilal Mohsin, Najla Zabani, Nasser Odah, Fatmah yamani, Lama Hefni, Naief Alhowaiti, Afnan Al Mutairi, Muhammad Talha kausar, Nadeem Shafique Butt, Wael Habhab

## Abstract

**Background:** ABO-incompatible (ABOi) kidney transplantation is increasingly utilized to address donor shortages in end-stage kidney disease (ESKD) patients. However, the impact of immunosuppressive regimens on infection risks remains a concern. This study examines the spectrum of infections, associated risk factors, and their influence on graft outcomes over a 5-year period.

**Methods:** A retrospective analysis of 24 adult ABOi kidney transplant recipients (2015–2019) was conducted, with follow-up until December 2024. Desensitization included rituximab, plasma exchange (PLEX), and IV immunoglobulin (IVIG). Infections were classified as bacterial, viral, fungal, or other opportunistic infections and their associations with graft survival and rejection were assessed.

**Results:** A total of 49 infectious episodes were recorded in 19 patients (79.2%); 5 patients had infection free follow-up of plus 5 years. Urinary tract infections (UTIs) were most common (23/49), followed by COVID-19 (11/49) and Influenza A (7/49). No episode of fungal infection was observed. Infection incidence was highest in females (52.6%), diabetics (47.4%), and patients with prior rejection episodes (10.5%). Kaplan-Meier analysis showed significantly lower infection-free survival in patients with graft rejection (p=0.0086). Despite frequent infections, overall graft survival remained high (91.7%), with no direct statistical association between infections and rejection.

**Conclusion:** Infections are prevalent in ABOi kidney transplant recipients, particularly in high-risk subgroups including females, patients with diabetes and prior graft rejection. However, long-term graft survival remains favorable with no association between infections and graft rejection. Optimized immunosuppression and infection surveillance are crucial for improving patient outcomes. Larger multicenter studies are warranted to validate these findings.

## Introduction

Kidney transplantation is a critical therapeutic option for patients with end-stage kidney disease, offering improved quality of life and survival rates compared to dialysis^1^. Although the shortage of compatible donors remains a significant challenge, ABO-incompatible (ABOi) kidney transplantation has emerged as a viable solution to expand the donor pool^2^. Despite its potential benefits, ABOi kidney transplantation is associated with a higher risk of complications, particularly infections, which can impact patient and graft-associated outcomes^3^.

Infections are a significant cause of morbidity and mortality in kidney transplant recipients, primarily because immunosuppressive therapy, which is necessary for the prevention of graft rejection, increases susceptibility to bacterial, viral, fungal, and parasitic infections. In ABOi kidney transplant recipients, intensified desensitization protocols, which may involve rituximab, intravenous immunoglobulin, and plasmapheresis, further exacerbate the risk of infections.

Several studies have highlighted the increased incidence of infectious complications in ABOi renal transplant recipients. For instance, a study conducted in Switzerland found an overall high rate of infections in ABOi kidney transplants compared to ABO-compatible (ABOc) kidney transplants. Another study reported a significantly higher rate of viral infections, including cytomegalovirus and polyomavirus, among ABOi recipients. Such infections in ABOi kidney transplant recipients might lead to graft dysfunction and an increased risk of graft failure.

Graft dysfunction in ABOi renal transplant recipients can be attributed to several factors, including acute rejection, chronic allograft nephropathy, and infection-associated complications^1,11^. Chronic allograft nephropathy, characterized by interstitial fibrosis and tubular atrophy, is often exacerbated by recurrent infections^12^. Viral infection can provoke T-cell-mediated immune responses that trigger the immunological cascade, leading to graft rejection. Although infections pose a significant challenge, existing evidence suggests that long-term graft survival outcomes remain encouraging in ABOi kidney transplant recipients^13^.

In Saudi Arabia, the local population’s unique demographic and epidemiological characteristics, coupled with the high prevalence of ESRD, necessitate a thorough understanding of infection-associated complications and their impact on graft function.

This study aims to comprehensively analyze post-kidney transplant infective complications and their association, if any, with graft dysfunction in ABOi renal transplant recipients at a tertiary care center in Saudi Arabia. By examining the incidence, spectrum, and outcomes of these complications, we hope to contribute valuable insights into ABOi kidney transplantation and improve patient care in the region.

## Materials and Methods

### Study Design and Population

This retrospective cohort study, which was approved by the Institutional Review Board of KFSHRC Jeddah (IRB number 2020-48), reviewed 24 adult ABO-incompatible (ABOi) kidney transplants performed at our center (KFSHRC Jeddah) between October 31, 2015, and December 30, 2019, with follow-up until December 31, 2024. Exclusion criteria included pediatric ABOi transplants and cases with major surgical or vascular complications requiring surgical re-exploration. Patient confidentiality was maintained using coded identifiers accessible only to authorized team members.The requirement of informed consent from the participants was waived due to the retrospective nature of the study

### Data Collection

Patient data, including demographics, comorbid conditions, anti-ABO titers, desensitization protocols, graft function, and infectious and noninfectious complications, were collected from the medical records. Infectious complications were identified with microbiologic cultures, serology, or polymerase chain reaction. Graft loss was defined as return to dialysis, retransplantation, or death with a functioning graft. Graft rejection was histologically confirmed by ultrasound-guided biopsy using the Banff criteria^14^.

### Desensitization Protocol

Pre-transplant desensitization reduced anti-ABO antibody titers to ≤1:8 perioperatively. Rituximab (375 mg/m²) was given 2–3 weeks before desensitization, followed by plasma exchange (PLEX) and intravenous immunoglobulin (IVIg, 0.2 g/kg/session), with a cumulative dose of 1–2 g/kg until titers reached ≤1:16. PLEX dose was calculated via following formula Plasma Volume (L) = 0.065 × Weight (kg) × (1 - Hematocrit)^15^

### Transplant Procedure and Immunosuppression

All transplants used living donor grafts, with anastomosis times between 30 and 55 minutes. Induction immunosuppression involved Thymoglobulin (4.5–6 mg/kg) starting intraoperatively (1.5 mg/kg initial dose). Mycophenolate mofetil, methylprednisolone, and tacrolimus were initiated as per protocol. Methylprednisolone was given at 250 mg on day 0, 125 mg on day 1, tapered by 5 mg daily to 20 mg, and then reduced weekly to a maintenance dose of 5 mg daily. Prophylactic valganciclovir (450 mg/day) was prescribed for 90 days for CMV D+/R+ patients and 6 months for CMV D+/R− patients. Cotrimoxazole (80 mg/day) was given for 6–9 months for PCP prophylaxis, initiated when eGFR exceeded 30 mL/min/1.73 m².

### Histological Assessment

Renal biopsies were performed when serum creatinine failed to improve within two weeks post-transplant, or for rises >27 μmol/L, proteinuria, hematuria, or suspected rejection. Protocol biopsies were not performed due to patient refusal.

### Outcomes of Interest

Primary outcomes were infections, including UTIs, viral infections (COVID-19, BK virus, CMV), fungal infections, and bacteremia. Secondary outcomes included graft rejection (antibody-mediated or T-cell-mediated per Banff criteria), graft failure, and mortality.

### Statistical Analysis

Data were analyzed using SPSS. Continuous variables were expressed as means ± standard deviations (SD) or medians with interquartile ranges (IQR). Categorical variables were presented as frequencies. Fisher’s exact test assessed associations between predictors and outcomes.

Kaplan-Meier survival curves were used to evaluate event-free survival from graft failure. A p-value ≤0.05 was considered statistically significant.

## Results

Table 1 outlines the demographic and clinical parameters of our study population.

**Table 1.**
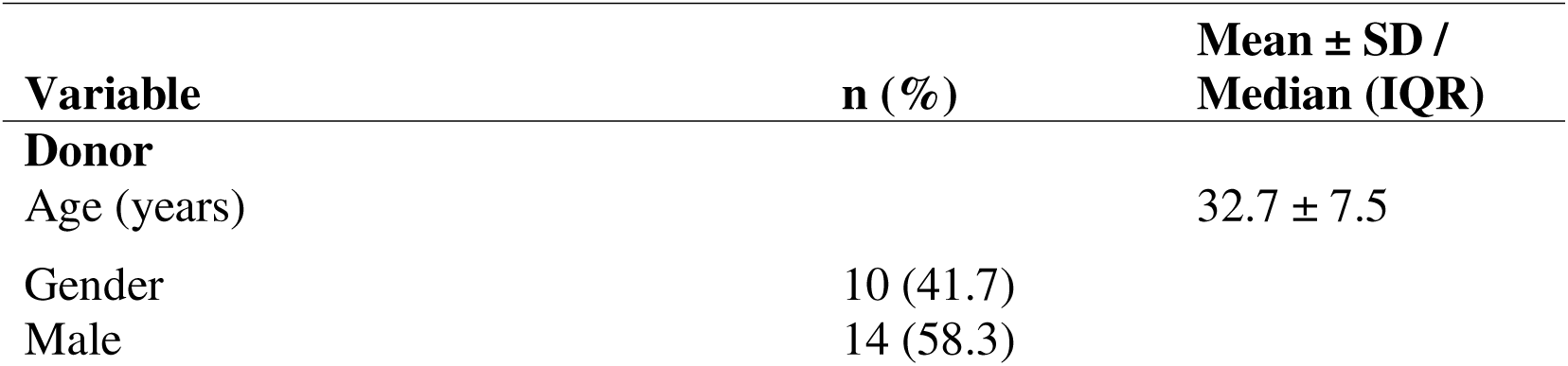
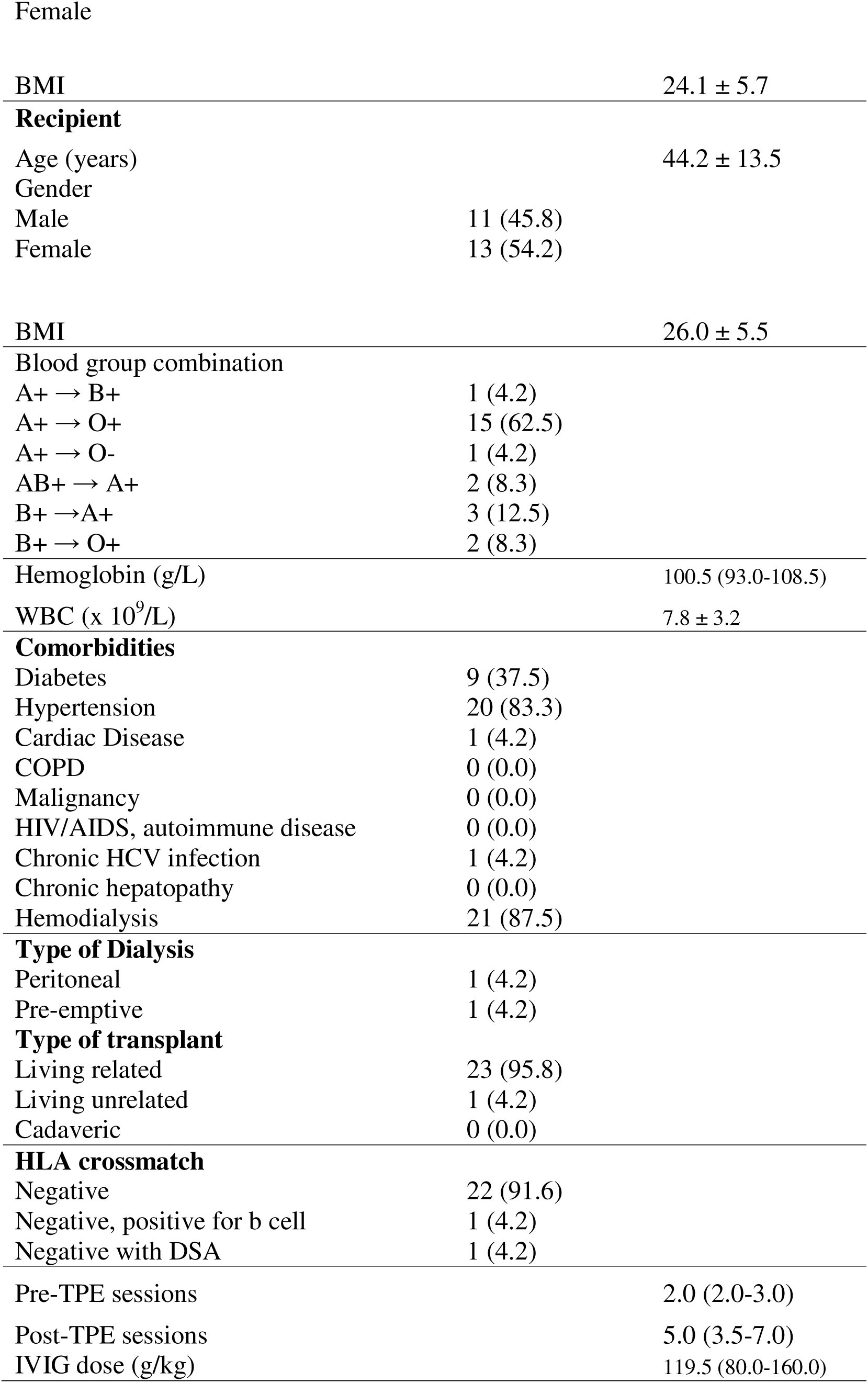
Demographic and clinical characteristics of ABO incompatible kidney transplant patients (n=24)

| Variable | n (%) | Mean ± SD /<br>Median (IQR) |
| --- | --- | --- |
| **Donor** |  |  |
| Age (years) | | 32.7 ± 7.5 |
| Gender | 10 (41.7) |  |
| Male | 14 (58.3) |  |
| Female |  |  |
| BMI |  | 24.1 ± 5.7 |
| **Recipient** |  |  |
| Age (years) |  | 44.2 ± 13.5 |
| Gender |  |  |
| Male | 11 (45.8) |  |
| Female | 13 (54.2) |  |
| BMI |  | 26.0 ± 5.5 |
| **Blood group combination** |  |  |
| A+ → B+ | 1 (4.2) |  |
| A+ → O+ | 15 (62.5) |  |
| A+ → O- | 1 (4.2) |  |
| AB+ → A+ | 2 (8.3) |  |
| B+ → A+ | 3 (12.5) |  |
| B+ → O+ | 2 (8.3) |  |
| Hemoglobin (g/L) |  | 100.5 (93.0-108.5) |
| WBC (x 10<sup>9</sup>/L) |  | 7.8 ± 3.2 |
| **Comorbidities** |  |  |
| Diabetes | 9 (37.5) |  |
| Hypertension | 20 (83.3) |  |
| Cardiac Disease | 1 (4.2) |  |
| COPD | 0 (0.0) |  |
| Malignancy | 0 (0.0) |  |
| HIV/AIDS, autoimmune disease | 0 (0.0) |  |
| Chronic HCV infection | 1 (4.2) |  |
| Chronic hepatopathy | 0 (0.0) |  |
| Hemodialysis | 21 (87.5) |  |
| **Type of Dialysis** |  |  |
| Peritoneal | 1 (4.2) |  |
| Pre-emptive | 1 (4.2) |  |
| **Type of transplant** |  |  |
| Living related | 23 (95.8) |  |
| Living unrelated | 1 (4.2) |  |
| Cadaveric | 0 (0.0) |  |
| **HLA crossmatch** |  |  |
| Negative | 22 (91.6) |  |
| Negative, positive for b cell | 1 (4.2) |  |
| Negative with DSA | 1 (4.2) |  |
| Pre-TPE sessions |  | 2.0 (2.0-3.0) |
| Post-TPE sessions |  | 5.0 (3.5-7.0) |
| IVIG dose (g/kg) |  | 119.5 (80.0-160.0) |

WBC, white blood cells; COPD, chronic obstructive pulmonary disease; DSA, donor-specific antibodies; Mean ± SD or Median (IQR) reported according to distribution of the variable data

WBC, white blood cells; COPD, chronic obstructive pulmonary disease; DSA, donor-specific antibodies; Mean ± SD or Median (IQR) reported according to distribution of the variable data

A total of 49 episodes of infections were observed in 19 patients, with 5 patients having infection-free follow-up over 5 years. 59%(n=29) episodes of infections occurred within the first 12 months of transplant while 77.5 % (n=38) infection were encountered within the first 18 months after renal transplant. There were 28 episodes of bacterial infections and 21 episodes of viral infections. There was no episode of fungal infection. The mean age of patients experiencing infections is 46 +/-11.1 years, with a median of 51.2 years. 10 of 19 (52.6%) patients were females, and 9 were males. All of these individuals were first kidney transplants. Nine patients had diabetes, while 13 were hypertensive. Two of them had rejection needing intensifying immunosuppressives prior to infective episodes. No patient experienced infections prior to graft rejection.

Thirteen patients had infective episodes with both bacterial and viral infection whether separately or together while six patients had either bacterial or viral infective episode. For the patient with, both, bacterial and viral infections, the mean age 52.3 +/-4.1 years. Nine out of thirteen such patients were females, 7 were having diabetes, and two had received treatment for graft rejection in the past.

Table 2 shows the distribution of infective episodes in the transplant patients.

**Table 2.**

|  |  |
| --- | --- |
| CMV | 2 |
| COVID | 11 |
| Bk Virus | 1 |
| Influenza | 7 |
| UTI | 23 |
| Bacteremia | 5 |
| Fungal infection | 0 |

UTI was the most common infection encountered in the study population, with 23 episodes of infection in 15 patients.(Figure.1). Most of the UTI’s (19 episodes) happened in first 12 months post kidney transplant. The mean age of patients having UTI was 49 +/-3.2 years. Female patients encountered UTI more frequently (17 out of 23 encounters) than males. Five patients had recurrent UTI: 4 females and one male; all 5 of them had diabetes. The most commonly encountered microorganisms were ESBL EColi(n=11), MDR Klebsiella (n=8), and Enterobacter (n=3). On five occasions, the patient developed bacteremia, needing parenteral antibiotics for more than one week.

**Figure 1.**
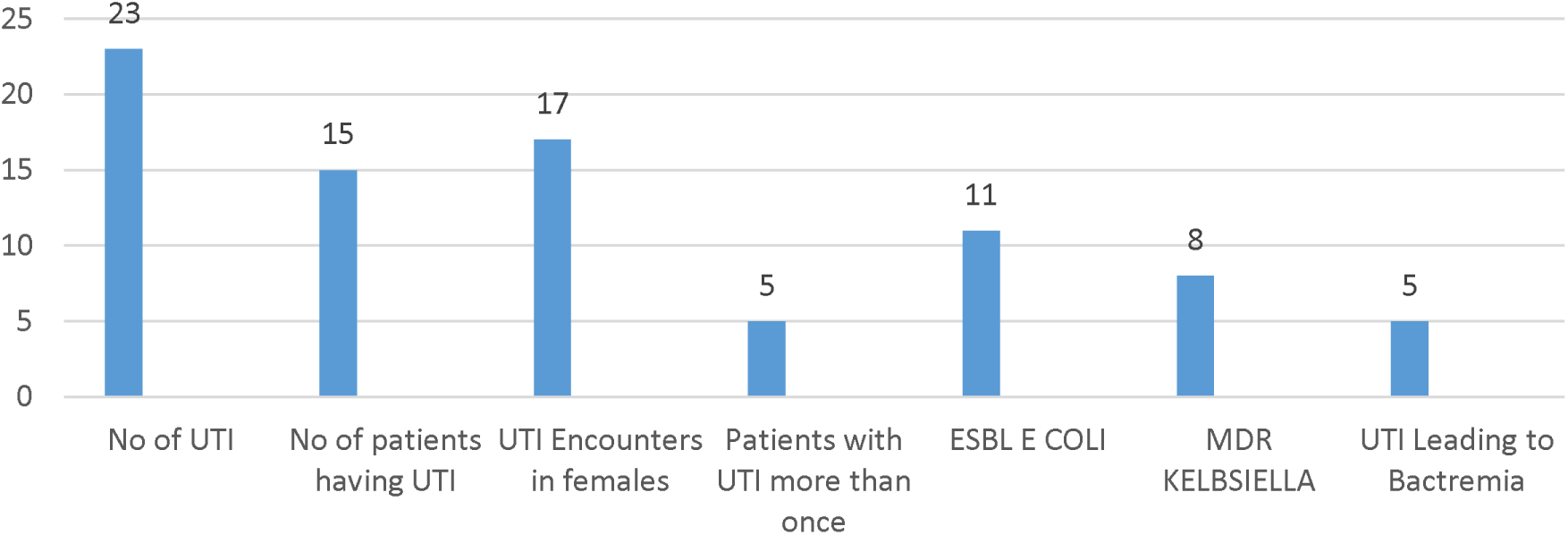
Summary of UTI in Patient Population

We encountered COVID-19 infection in eight patients (n=8), and the total number of encounters with COVID-19 infection was 11, with three patients getting infected twice with COVID-19 infection. Five out of eight patients (62.8%) were females, and six (75%) had diabetes. Two of these patients received treatment for graft rejection in the past 6 months. There was no episode of graft loss or mortality during their infective episode with COVID. Five patients had COVID-19 category B, and 3 had Covid category C symptoms. Two out of 3 patients with COVID category C needed Intensive care monitoring and treatment. Patients needing ICU care developed transient AKI that resolved within 2 weeks of follow-up. As our centre protocol, we discontinued MMF for patients with COVID-19 category B and C with reduction of target Prograf levels to 3-4 ng/ml with dose increase of prednisolone to 10 mg daily. MMF was resumed one week after complete resolution of symptoms. Influenza was encountered in seven patients. Patients needed hospitalization for 3-5 days and received Osaltamiver for 5 days, followed by complete recovery. We experienced CMV viremia in 8.3 % (n=2) individuals. One patient was diagnosed incidentally on routine q 6 months screening, and the other had GI symptoms needing a workup that revealed CMV viremia. Both incidences occurred between 6 to 18 months after the renal transplant, and both patients had diabetes. We observed BK viremia in one patient who was diagnosed while working up a rising renal profile in the patient. BK virus titers were more than one million copies/ml. Despite modification of immunosuppression, the patient with BK nephropathy progressed to develop CKD with a functioning graft at the date of the last followup (55 months).

We did not experience fungal infections, malignancy, or mortality in this group.

The Kaplan-Meier plot reveals a clear difference in infection-free survival between individuals with and without rejection.(Figure.2) The group without graft rejection (“Graft rejection=No”) appears to maintain a higher probability of being infection-free over time compared to the group with graft rejection (“Graft rejection=Yes”).

**Figure 2:**
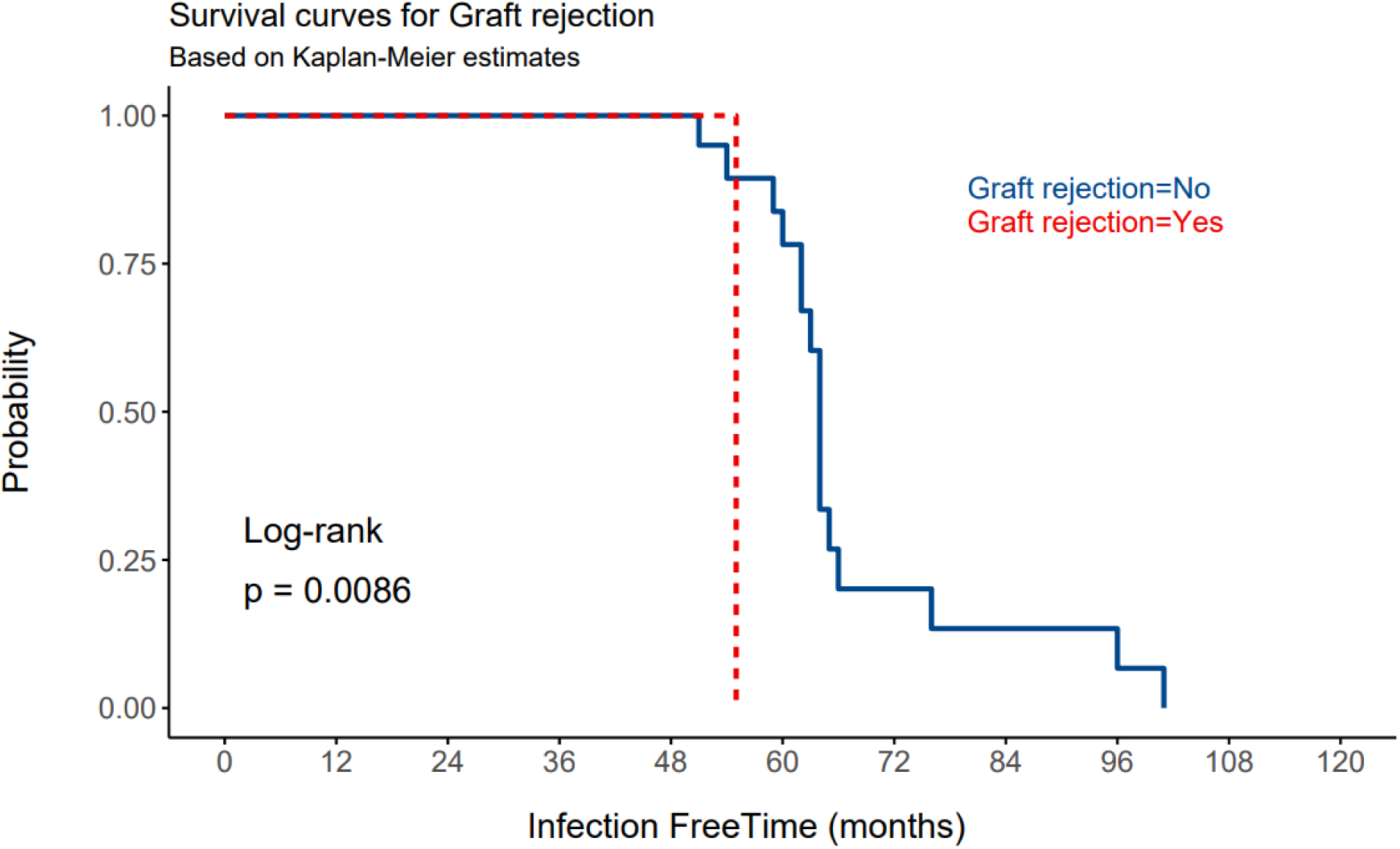
Infection-Free Survival by Graft Rejection Status: Kaplan-Meier Analysis with Log-Rank Test

The Log-Rank test yields a p-value of 0.0086, less than the conventional threshold of 0.05. This indicates a statistically significant difference in infection-free survival between individuals with and without graft rejection. The plot indicates that the probability of staying infection-free is consistently higher for the group without graft rejection. It also shows higher frequency and clustering of infective events in the early post-kidney transplant period.

Our chart review suggests that female gender, diabetes mellitus, and prior history of rejection significantly increase the risk of infections in the renal transplant population.

## Discussion

In the present study, bacterial and viral infections were common in patients undergoing ABOi kidney transplantation. These infections were more frequent in female patients, patients with diabetes, and patients who experienced graft rejection requiring intensified immunosuppressive therapy. Notably, most of the infections occurred during the first 18 months after kidney transplantation. UTIs were the most frequent infections, followed by COVID-19, influenza A infection, bacteremia, and BK nephropathy. However, the graft survival rate was 91.7% during a mean follow-up period of 64 ± 12.6 months despite these complications. Severe antibody-mediated rejection was the cause in both patients who experienced graft failure during the post-transplantation follow-up. However, a direct association between infections and long-term graft dysfunction was not observed, supporting the current evidence indicating that infection-related complications can be managed without compromising graft survival^16,17^

Our study reveals that bacterial and viral infections are commonly observed in ABO-incompatible (ABOi) renal transplant patients. They are more frequently observed in females, patient with diabetes, and patients who have experienced graft rejection needing intensified immunosuppressive therapy. The infective episodes have been observed to be significantly clustered within first 18 months post kidney transplant. Specifically, urinary tract infections (UTIs) were the most frequent, followed by COVID-19, Influenza A, bacteremia, and BK nephropathy (4.2%). Despite these complications, we observed encouraging graft survival of 91.7% over a mean follow-up period of 64 ± 12.6 months. We noted two cases of graft failure (8.3%) due to severe antibody-mediated rejection (ABMR). However, no direct correlation was found between infections and long-term graft dysfunction, supporting existing literature that infection-related complications can be managed without compromising graft survival^16,17^.

A locoregional study in Saudi Arabia reported similar trends in infection rates among kidney transplant recipients, highlighting the prevalence of bacterial infections such as *E. coli* and *Klebsiella pneumoniae*^18^. These infections were also considered major contributors to graft dysfunction in a Saudi cohort, a finding not consistent with our observations^19^. This discrepancy may stem from the higher immunologic risk in Saudi cohorts, which may underlie the development of graft dysfunction during the peri-rejection period, rather than infections. Another study in Saudi Arabia also reported that the rate of infectious complications was higher in ABOi kidney transplant recipients than in ABO-compatible kidney transplant recipients and that UTIs were the most common type of infection^20^, in agreement with our findings in the current cohort.

Our findings align with international studies reporting increased infection risk in ABOi kidney transplant recipients due to intensified desensitization protocols^7,9^. The notable absence of fungal infections in our cohort is in contrast with global studies reporting fungal complications in 5%–10% of the kidney transplant recipients^10^; our observation might be attributed to stringent prophylactic measures in our centre and regional environmental and population-based immunologic factors; however, the possibility of under-diagnosis due to limited routine fungal screening and the impact of the small cohort size cannot be ruled out.

A study from Switzerland also reported higher infection rates in ABOi kidney transplant recipients compared with ABO-compatible kidney transplant recipients, with a particular emphasis on viral infections, such as those caused by CMV and polyomavirus^19^. In the present study, we also observed a significant rate of viral infections, including COVID-19 and influenza A infection, contributing to the complexity of managing these patients. Moreover, although the infection trends in our cohort were comparable to that reported in studies from Europe and North America, we observed relatively lower rates of CMV and BK viremia^21^. This discrepancy may reflect differences in preemptive antiviral strategies and population-based immunologic responses, although the small cohort size might have also contributed to the observed difference.

Overall, our findings are consistent with both locoregional and international studies, indicating a higher susceptibility of infections in ABOi kidney transplant recipients, indicating that the detection and targeted management of infections are essential for minimizing complications, especially in high-risk groups such as female patients, patients with diabetes, and patients with graft rejection. Despite the high prevalence of infectious complications, we observed encouraging graft outcomes in ABOi kidney transplant recipients, which emphasizes that higher risk of infectious complications should not be a hindrance in pursuing ABOi kidney transplantation, because of the encouraging long-term graft outcomes.

## Limitations

Despite its valuable insights, our study has several limitations. The small cohort size restricted the statistical power to detect subtle associations between infections and graft outcomes. Due to the single-center study design, the study results may not be generalizable to diverse populations with varying immunosuppressive practices. Moreover, the absence of protocol biopsies might have led to the underreporting of subclinical rejection episodes. Finally, we acknowledge the potential selection bias due to the retrospective data collection and the concern regarding potentially missing information on infections that might have been diagnosed or treated outside the study institution.

## Conclusion

This study highlights a higher incidence of infections in subgroups of ABOi kidney transplant recipients including females, patients with diabetes and prior history of rejection. However, despite infective complications, the overall graft survival of 91.7%, over a follow-up of plus five years, is very encouraging. There was no statistical correlation between infections and graft dysfunction. Hence the concern of infective complications should not be considered a barrior for proceeding with ABOi kidney transplant.. We suggest a prospective large multicenter trial for better understanding the infective and graft associated outcomes in ABOi kidney transplant.

## Data Availability

All data produced in the present study are available upon reasonable request to the authors
All data produced in the present work are contained in the manuscript

